# Feasibility and acceptability of collecting dried blood spots (DBS) from children after vaccination during Supplementary Immunization Activities to estimate measles and rubella seroprevalence

**DOI:** 10.1101/2024.02.14.24302830

**Authors:** Andrea C. Carcelen, Christine Prosperi, Mutinta Hamahuwa, Kelvin Kapungu, Gershom Chongwe, Francis D. Mwansa, Phillimon Ndubani, Edgar Simulundu, Innocent C. Bwalya, Kalumbu H. Matakala, Gloria Musukwa, Irene Mutale, Evans Betha, Nchimunya Chaavwa, Lombe Kampamba, Japhet Matoba, Passwell Munachoonga, Webster Mufwambi, Ken Situtu, Philip E. Thuma, Constance Sakala, Princess Kayeye, Shaun A. Truelove, Amy K. Winter, Matthew J Ferrari, William J. Moss, Simon Mutembo

## Abstract

Nested serosurveys within routine service delivery platforms such as planned supplemental immunization activities (SIAs) provide an opportunity to collect information that can be used to answer valuable questions on the effectiveness and efficiency of the delivery model to inform future activities. However, integrating research data collection in SIAs is rarely done due to concerns it will negatively impact the program.

We conducted a serosurvey nested within the November 2020 measles-rubella SIA integrated with the Child Health Week activities in Zambia to evaluate this approach. In-depth interviews with the study teams and vaccination campaign staff at the vaccination sites were conducted. Recorded interviews were transcribed, transcripts were coded and then grouped into themes based on a process evaluation framework. A multi-methods analytical approach was used to assess the feasibility and acceptability of collecting dried blood spots from children during the SIA. This included a quantitative assessment of participant enrollment.

The serosurvey successfully enrolled 90% of children from Child Health Week due to close coordination and teamwork between the vaccination teams and serosurvey team, in addition to substantial social mobilization efforts. Continually adjusting the sampling interval that was used to select eligible children allowed us to enroll throughout the SIA and capture a representative sample of children in attendance although it was challenging for the staff involved.

As vaccination programs aim to tailor their approaches to reach the hardest-to-reach children, embedding research questions in SIAs will allow evaluation of the successes and challenges and compare alternative approaches. Lessons learned from this experience collecting data during an SIA can be applicable to future research activities embedded in SIAs or other delivery platforms.

## Introduction

Serosurveys for vaccine–preventable diseases (VPD) are cross-sectional surveys that measure antibodies against pathogens from a representative sample of a population to estimate immunity (1). Traditionally, vaccine coverage and disease surveillance data are used to infer immunity levels, but these sources do not directly measure immunity (2). Poor data quality can lead to an overestimation of immunity and population groups with immunity gaps may be missed by both routine and SIA vaccination activities. Serosurveys can be used to complement data from other sources, including vaccine coverage and disease surveillance. They are most relevant to predict the risk of outbreaks and plan for vaccination campaigns in the specific situation where longstanding immunity gaps are suspected but coverage data are not adequate to assess the risk because of quality issues, migration of populations, or gaps in specific age or other sub-groups in the population (3).

Serosurveys can use previously collected specimens from bio banks or health facilities for testing or prospectively collect specimens. Specimen collection may be done through purposively designed surveys or by nesting within other planned surveys, such as Demographic Health Surveys (DHS) or Post Campaign Coverage Surveys (PCCS) (4, 5). New specimen collection allows for better control of data collection and sampling methodologies but can be costly and logistically challenging (6, 7). Nesting within other planned surveys results in cost saving compared to a standalone serosurvey (7). However, there are still considerable personnel and transport costs to visit communities to conduct data and specimen collection, even in a nested design. This is especially relevant for surveys using multi-stage sampling which typically require multiple days to map and enumerate the community prior to enrollment (8, 9).

An alternative approach to save costs is to nest a survey within delivery platforms such as supplemental immunization activities (SIAs) where caregivers are already bringing their under five children to receive services. SIAs provide an opportunity to easily identify and collect data and specimens from children in the age of interest for many VPDs. This information can be used to answer valuable questions on the effectiveness and efficiency of the delivery platform to inform future activities. Integration of serosurveys and other research studies in the SIA platform has been avoided due to concerns that such research activities would negatively impact the success of the SIA. However, integration of other services in the SIA platform, such as bed net distribution, is routinely done and recommended by the World Health Organization as comprehensive approach to service delivery (9, 10).

Child Health Weeks (CHWs) are semi-annual, campaign-style, facility and outreach-based events that were initiated in Zambia in 2003 to provide a package of high-impact preventive services to under five children. These services included vitamin A supplementation, growth monitoring and promotion, vaccinations, deworming and promotion of intermittent treatment of insecticide treated mosquito nets (11). We nested a serosurvey in the November 2020 SIA to assess the seroprevalence of measles and rubella among children attending the 2020 SIA integrated in the Child Health Week. In this manuscript we evaluate the processes, feasibility, and acceptability of nesting a serosurvey in the SIA. We describe the planning and implementation of the serosurvey nested in the SIA, quantitative summaries of participant enrollment and survey operations, and qualitative summaries from in-depth interviews with data collectors, supervisors, and vaccinators. The vaccination coverage and seroprevalence findings are presented in a separate manuscript (12).

## Materials and Methods

We conducted a serosurvey nested within the November 2020 measles-rubella SIA integrated with the Child Health Week activities in Zambia which targeted children 9 to 59 months of age (12). We implemented this serosurvey in two districts in Zambia: Ndola District, Copperbelt Province (primarily urban), and Choma District, Southern Province (primarily rural). We included 15 vaccination sites in each district. At each site there were two teams: the vaccination team and the serosurvey study team. When a caregiver arrived at the site, they went to the vaccination table. There was a study staff stationed near the vaccination team who systematically sampled children as they left the vaccination table. The selected child and their caregiver were then directed to the survey team table. If the caregiver consented to take part in the serosurvey, a questionnaire was administered, and a Dried Blood Spot (DBS) sample was collected from the child by finger prick onto a well-labeled card. The sample was then dried, stored, and transported to the central laboratory for testing. Additional details regarding implementation, sampling and tracking are provided in the Supplemental Methods (S1 Appendix).

We used a multi-methods analytical approach to assess the feasibility and acceptability of collecting dried blood spots from children at the point of vaccination during a vaccination campaign. This included a quantitative assessment of how many children were expected to attend, how many ended up being eligible and how many were enrolled and provided specimens. To evaluate the process of integrating specimen collection during the campaign, we conducted in-depth interviews with both the study teams and vaccination campaign staff at the vaccination sites where DBS was collected.

All study staff and vaccination campaign staff at sites where specimens were collected were eligible to be interviewed. Purposive sampling was used to identify respondents. Co-investigators at Macha Research Trust and Tropical Diseases Research Centre conducted data collection. Interviews were conducted using a semi-structured interview guide in English and were audio recorded and transcribed. Recruitment was based on availability and interest of staff and included all 3 study supervisors, 10 of the 30 study team staff and 10 of the 30 vaccination campaign staff. All data collection occurred at the end of the day after campaign activities were completed or in the days immediately following the campaign. The recruitment period for the interviews was from November 23 to December 8, 2020. Forty-two respondents were interviewed between the two districts, including 18 serosurvey staff, 6 serosurvey supervisors, and 18 SIA vaccinators (S1 Table).

All transcripts were coded initially by one research team member in Dedoose v9.4, and 20% of transcripts were double coded by the principal investigator. A codebook was developed based on previous acceptability of serosurvey work conducted in Zambia but was updated with additional codes as needed Disagreements were resolved by consensus. Initial codes were grouped into axial codes and subsequently themes based on programmatic aspects addressed using a process evaluation framework We used constant comparison to assess any differences by study team member or vaccinator and barriers or facilitators to the program.

Ethical approval was obtained from the Tropical Diseases Research Centre ethics review committee, and Johns Hopkins Bloomberg School of Public Health Institutional Review Board. Further regulatory approval was given by the Zambia National Health Research Authority. Written consent was obtained from caregivers before enrolment in the serosurvey, and for in-depth interviews of health workers verbal consent was obtained from serosurvey staff and vaccinators.

## Results

Below we present details on the steps followed to operationalize the serosurvey nested in the SIA, including a qualitative evaluation of the serosurvey planning, staffing and training, social mobilization, enrollment, sampling, and implementation. Quantitative summaries of sampling and serosurvey enrollment contextualize the qualitative findings on perceptions.

### Stakeholder engagement and planning

During the planning phase of the serosurvey we used the district SIA microplans and input from district health management teams to estimate the number of children to be vaccinated in each fixed and outreach vaccination site. We inflated the target number of children to be vaccinated at each serosurvey location by 5% then estimated the number to be vaccinated on each of the five or six days of the SIA. We planned to enroll between 70-85 children per survey location, aiming for 10-15 children per day to distribute enrollment across all days of the SIA. We set a daily maximum to prevent over enrolling on a given day if the volume of children was larger than anticipated. Using the estimated number of children to be vaccinated per day and the overall and daily target enrollment numbers we proposed an initial sampling interval for each location.

Respondents highlighted the importance of engaging with Ministry of Health through the district health office, specifically Maternal and Child Health staff, from planning and training through implementation. Engagement at these higher levels was critical to facilitate engagement with staff at the selected health facilities, as noted by a supervisor.

> *“So we had to work with the district health office, we worked with the matron who supervises the mother to child health unit and they put us in touch with the sisters in charge for every site that was selected through the DHO, then that is where the teams were reconstituted” -T_SUPER_1*

### Staffing and training

The nurses in-charge at the selected health facilities played a key role in the survey by identifying staff to conduct the serosurvey without compromising the needs of Child Health Week, which required rearranging staff schedules or requesting that staff on leave return to work. Staff were experienced nurses, including some with prior survey experience, who were familiar with the community where the survey was conducted, which was a substantial strength for the survey. All staff were trained on all activities, such as obtaining informed consent, entering data in the tablet, and collecting dried blood spots. Staff noted they felt equipped to address questions from caregivers during the SIA, and appreciated the experience gained with regards to social mobilization and speaking to caregivers, conducting research, and the collection of dried blood spots. However, some serosurvey staff and supervisors noted the training period was insufficient and too fast-paced. Supervisors leading training sessions also noted additional time was needed to review materials in advance of the training. Prior to the SIA, serosurvey staff conducted a pilot exercise at health facilities where they practiced sampling children, introducing the survey, obtaining consent, and conducting the interviews with families attending routine immunization (RI). While many highlighted the value of the piloting exercise, some also noted additional piloting experiences were needed. Both serosurvey staff and vaccinators mentioned that including the vaccinators in the training would have been valuable to make them aware of the survey activities and motivate them to support the survey staff in sensitizing parents leading up to and during Child Health Week. This was reported to have happened in some sites, but not others. As one supervisor noted, staff should include *“incorporating a lot of people, community health workers as well, because those help very much in social mobilization. Imagine we bought a bicycle for each one of them, social mobilization was going to be very easy.”-T_SUPER_3*

### Community mobilization

Prior to field work, the survey team worked closely with the maternal and child health (MCH) coordinators and health promotion officers at the district office to conduct community sensitization for the serosurvey, which in many locations was closely linked to the sensitization for the SIA. Staff said this included home visitations, megaphone, and radio advertisement, distribution of information, education, and communications (IEC) materials such as leaflets and posters at clinics, and in-person meetings held at churches to inform people about the upcoming serosurvey. However, during data collection, staff mentioned that in some areas sensitization was done well while in others it was insufficient. Staff reported that some parents or caregivers only learned about the serosurvey when they visited the health facility during child health week. Throughout the week, the community health workers and health promotion officers intensified community outreach activities due to low turnout. Having health facility staff actively involved in sensitizing parents or caregivers who brought their children for routine immunization, made parents receptive to the serosurvey. Some staff recommended that community sensitization should be done earlier so that people are well informed about the survey to avoid misconceptions, as noted by a serosurvey data collector.

> *“Maybe just prolonging the period for community sensitization, so that facilities are with the information for a longer period so that as they talk to the community members who are coming for each and every service be it out patient department or during the maternal and child health clinics they are able to talk to them” -MK-Sero-1*

### Enrollment

In total 2,942 children attending the campaign at the selected locations were approached by the survey team, out of an approximate 25,088 children vaccinated. Ninety percent of children approached at campaign sites in Ndola District were enrolled, but there was lower participation in Choma District (74% (Fig 1). Refusals in Choma District were concentrated in three campaign sites, where participation ranged from 31% to 61%. Approximately three to four percent of children approached in each district could not be enrolled because no caregiver was available to provide consent. While the daily enrollment target at a site was intended to be 10-15 children, the number enrolled on a given day ranged from 2 to 49 (S1 Fig), varying as we adjusted the daily maximum.

**Fig 1.**
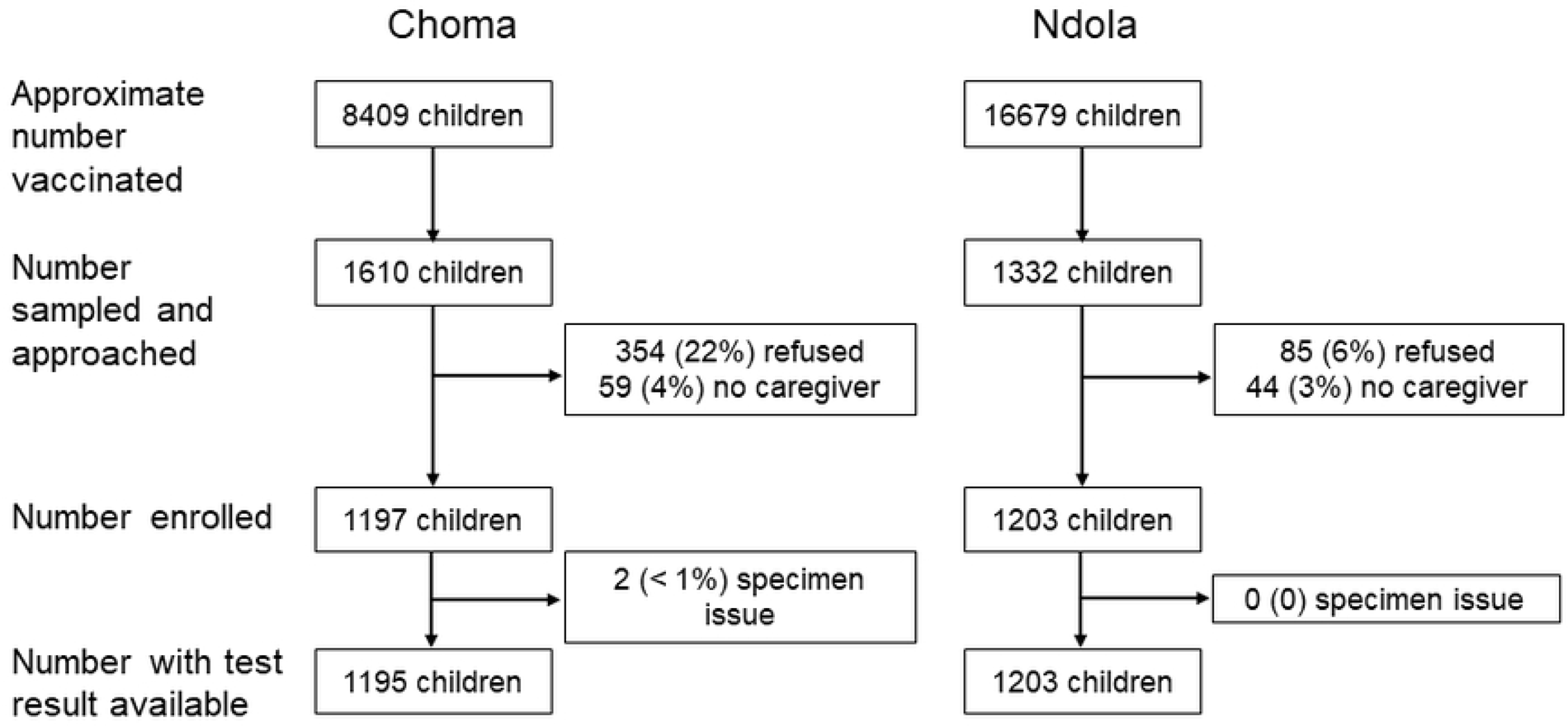
Participant cascade by district at selected health facilities where serosurvey was conducted. Number of children vaccinated at the select serosurvey locations was estimated based on serosurvey staff observations and discussions with the health facility staff.

Survey staff recorded the number of children vaccinated each day based on their observations as they were sampling children or from the nurses in-charge. We summarized the number of children vaccinated at each survey location and compared it to the number expected based on the microplan and communications with health facilities during the planning phase (Fig 2). In Choma District, we observed fewer children vaccinated than we had expected in 12 of the 15 campaign sites where the serosurvey was conducted, including 6 with fewer than half of what was anticipated. The opposite occurred in Ndola District, where 12 of the 15 campaign sites had more children vaccinated than expected, including four with more than double what was anticipated. Overall, we observed approximately 35% fewer children and 22% more children vaccinated in Choma and Ndola districts, respectively, compared to expected based on the information used in planning. This discordance in the number expected versus observed resulted in shifts to the sampling interval to ensure we met the overall targets and distributed enrollments throughout the SIA, without overburdening the survey staff on any given day.

**Fig 2.**
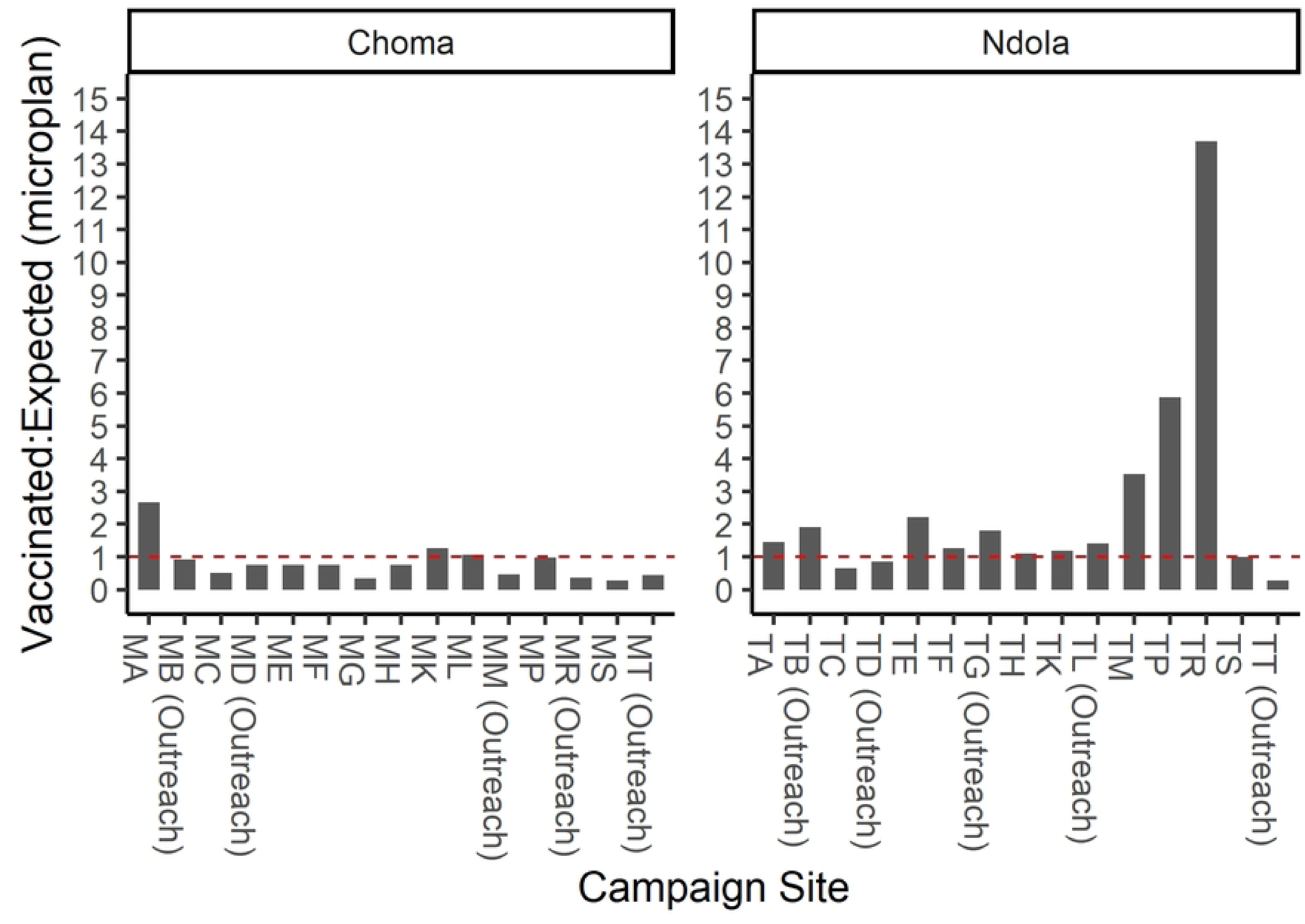
Ratio of number of children vaccinated to number expected by campaign site. Number of children expected to be vaccinated at a given campaign site was obtained from the microplans with input from local research and health facility staff. Range for number of children expected per site: Ndola District 99-2828; Choma District 110-2193. Range for number of children vaccinated per site: Ndola District: 324-1924; Choma District: 158-1004.

### Sampling interval

While we assumed the same number of children vaccinated per day for planning purposes, we observed the volume of children varied day to day. Therefore, the sampling intervals were changed during the campaign, ranging from every child to every 30th child, depending on the facility and day (S2 Fig). Nine of the thirty facilities used the same sampling interval on all days. Five facilities began with a very high sampling interval (e.g., every 20th child) then needed to reduce on subsequent days to increase the numbers enrolled. Other facilities had less extreme but more frequent changes throughout the week. Each evening after teams uploaded data and reported progress on WhatsApp, the central study team reviewed and proposed changes to the sampling interval based on input from the supervisors and survey teams.

Changes were made based on how many children had been enrolled that day and reasons for differences in observed enrollment compared to planned enrollment, expected attendance in the coming days at a given SIA site, and study approved sample size and the by-site enrollment goals to avoid any issues with considerable under or over-enrollment overall or at a given site and maintaining a balance between fixed and outreach sites. Operational factors that affected observed enrollment included the number of serosurvey staff relative to the volume of children; issues or events that impacted the volume of children attending the SIA (e.g., weather, when routine immunization services are usually provided, visits by dignitaries), or issues impacting ability of the serosurvey staff to conduct enrollment (e.g., lack of transport, SIA site closed for the day or opened late, SIA site extending vaccination activities in Choma District). Changes to the sampling interval were communicated to the site coordinators who disseminated it to the survey staff, however due to the time required to review and propose changes the survey teams typically were not notified of changes until the following morning as they were beginning that day’s work.

### Perceptions of the sampling interval

During interviews survey staff described how they implemented the systematic sampling procedures. Descriptions of how they communicated with caregivers about the sampling procedures included mentioning the survey is conducted on a subset of the population representing others and that a system is in place to select those children. Some referred to the process as based on ‘luck’ or being ‘done randomly’, rather than systematic sampling. There were also a few deviations from standard practice described in the interviews, such as skipping certain children who were sampled if the staff perceived that the caregiver would refuse or would not be able to provide consent for the child (e.g., maid or grandparent). Survey staff mentioned questions they received from caregivers related to selection, such as why only one sibling was selected. One supervisor described below how explanations were provided.

> *“We are just sampling, your child is representing part of the community around so it’s not that we are going to get blood from each and every child no, but what we are going to …the results that we are going to get from each child will have a representation of this same area or community” -T_SUPER_2*

Many staff noted challenges they faced during the survey due to the discrepancy between the volume of children observed relative to what was expected and was used to inform the sampling interval. There was mention of some outreach locations being overflowed and locations with low numbers on initial days which ramped up as additional sensitization occurred. Changes to the sampling intervals made by the central team led to confusion for the staff on the ground who needed to adapt procedures. It also led to confusion for community members who learned about the survey and every nth child being selected from a neighbor who attended Child Health Week on a prior day. One serosurvey staff member suggested autonomy in terms of selecting the sampling interval based on the daily situation since it may change day to day and even within a day. Despite these challenges, staff generally reported it was easy to meet their enrollment targets.

### Facilitators and barriers to enrollment

#### Perceived benefits

Survey staff used different ways to describe the value of the serosurvey and community-level benefits to caregivers. Many highlighted how the findings will be valuable to the community, stating that the study was being conducted to learn how children develop antibodies against measles and to ensure children in that area are fully protected. Most only mentioned measles in their descriptions, but a handful also mentioned rubella. Some noted how these findings would be used to guide future interventions, such as repeating vaccinations, targeting certain areas with lower immunity or investigating if there were issues with vaccine storage. Some staff members incorrectly described the serosurvey as informing development of a stronger vaccine. The descriptions of community-level benefits were generally positive; however, one staff member encountered a caregiver who was accepting of vaccines but began to question the vaccine in response to how the benefits were described. As noted by one serosurvey data collector, *We want to see how efficient the vaccines are and see what the government can work on to ensure that the children are fully vaccinated.” -TH_SERO_1*

In terms of individual-level benefits, staff discussed how caregivers appreciated the refreshments, and masks to a lesser degree. Some staff mentioned that caregivers at the campaign site who were not selected asked to participate to receive these incentives. Staff described how this led to frustration from caregivers whose children were not selected to participate. Some staff mentioned they asked participants not to tell others about the incentives to avoid issues and overcrowding. There were also concerns raised about the incentives representing a payment for the blood leading to suspicions. One vaccinator described how the benefits were well-received: *“When a child has recruited in the system there is a packet of sugar which is given to appreciate the mother, I think that way it has really made the whole process very good and I have seen even the mother goes out smiling.” -MH_Vac_1*

A common issue raised by participants was caregivers’ expectation to receive their child’s test result. Staff reported explaining that test results will not be provided but the findings will be summarized at the facility-and district-levels to benefit the broader community. One supervisor correctly highlighted how the test results only reflect immunity from prior vaccinations and the child’s immunity will change due to the vaccine they received during Child Health Week.

> *“You know people when you are doing a test they expect a result, so they were asking; “when are we getting the result?” but then when you explain that the result won’t really affect you, we want to see if the injections that were given earlier on has built the immunity with the kid, this injection that they gave today will only build immunity uhmmm weeks from now and it won’t change any… you know anything with the child, the result is not necessary at this point. They accepted it.” -T_SUPER_1*

### Misconceptions and reasons for refusal

Respondents cited several reasons why caregivers refused to participate in the serosurvey. For example, some caregivers were concerned about pricking the child again after they had just received the vaccine or reported a lack of time. There were also several misconceptions, mostly related to blood collection and confusion about the survey and its purpose. These included concerns the survey staff were witches or Satanists, and the blood was being used as a sacrifice for rituals. Staff believed that since the community is familiar with finger prick blood collection for HIV and malaria testing, caregivers questioned if their children were being selected for the serosurvey due to perceived illness of the child. One participant reported that finger/ thumb printing the consent form, used in place of a signature for illiterate caregivers, was a cult initiation. The survey was also mistakenly linked to a political party due to community members misinterpreting a label on supplies. Staff revealed that these misconceptions were spread by caregivers who attended the SIA then misinformed other community members, as a result, some potential participants refused to participate. As described by a serosurvey data collector, community members were also concerned that the blood was being tested for COVID-19.

> *“And went to explain to the other mothers that the blood they are collecting on those cards is for Covid so because of that, the community hearing about Covid they all shunned away saying you want to test us for Covid.” -TL-SERRO-1*

### Interpersonal influences

Staff also discussed how male partner involvement could be a motivator if the partner was supportive, or a barrier if mothers could not agree to participate in the survey without permission from their husbands. One participant described a situation where a mother enrolled a child in the serosurvey without consent from her husband, and he chased her from their home. Participants noted that sensitizing the community well in advance of the serosurvey would have allowed women time to consult with their husbands prior to the SIA.

### Implementation of the serosurvey

#### Supervision and logistics

The vaccination and survey staff felt that the survey was well organized. There was a supervisor at each site on the first day; and daily supervision at fixed and outreach sites was facilitated by supervisors having designated vehicles. There was also constant communication through the phone to answer any questions from the teams. The supervisors were also in constant communication with study principal investigators (PIs) and central laboratory teams, providing brief daily updates on each site. Summary reports were prepared by the central team to rapidly review data and provide feedback (S1 Appendix). Study staff mentioned there were no stock outs of study supplies, and there was efficient specimen transportation to the central laboratory.

Although the respondents felt the survey was a success there were a few challenges. For instance, some outreach sites did not have shelter in case of rain. There were a few difficulties in collecting blood and this led to having to do a second prick, children running away, fighting, fidgeting, crying, or having small fingers.

#### Coordination

Generally, the vaccination and serosurvey staff described working well together throughout the campaign, often mentioning that they worked “hand in hand”. A couple of staff members mentioned some difficulties on the first day due to logistical issues but were able to smooth these out thereafter. There were also some limitations noted by staff who were involved in vaccination. They explained that they learned of the survey when it was too close to implementation date, therefore they were denied an opportunity to participate in the planning meetings. A few vaccination staff felt that there was more work to be done during the current child health week compared to the past ones.

Overall, respondents agreed that nesting the serosurvey in the child health week was a success. Coordination between the vaccination and serosurvey staff was commonly mentioned by both as key to that success. Recruiting children attending the campaign was viewed as a good entry point, since other services are often provided there, and no detrimental effects on the vaccination campaign were mentioned. They also pointed to mothers’ willingness to bring and enroll their children, demonstrating the success of the program. One vaccinator mentioned there was no negative effect:

> *“I: what effect do you think the serosurvey had on vaccination campaign?*

> *R: “…there was no any negative effect at all, all was excellent, the whole process the whole program is excellent” -MH_Vac_1*

## Discussion

We successfully integrated a serosurvey into Child Health Week, enrolling 82% of children invited to participate. As highlighted by the qualitative interviews, much of the success was attributable to close coordination and teamwork between the vaccination teams and serosurvey teams. This began during the planning phase by working with the district health offices in the two districts and health facilities staff. Engaging local staff for the serosurvey who were already familiar with the community facilitated close coordination for implementation. We believe this heavy level of involvement of the Ministry of Health allowed for smooth integration of a research project into service delivery and should be the standard for research using a programmatic platform. In this paper we summarized the lessons learned of collecting dried blood spots from children attending an integrated Child Health Week based on the qualitative interviews (Table 1).

**Table 1.**
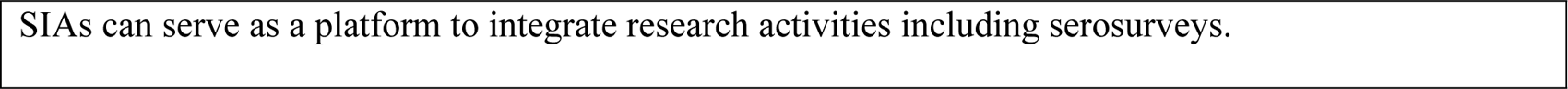
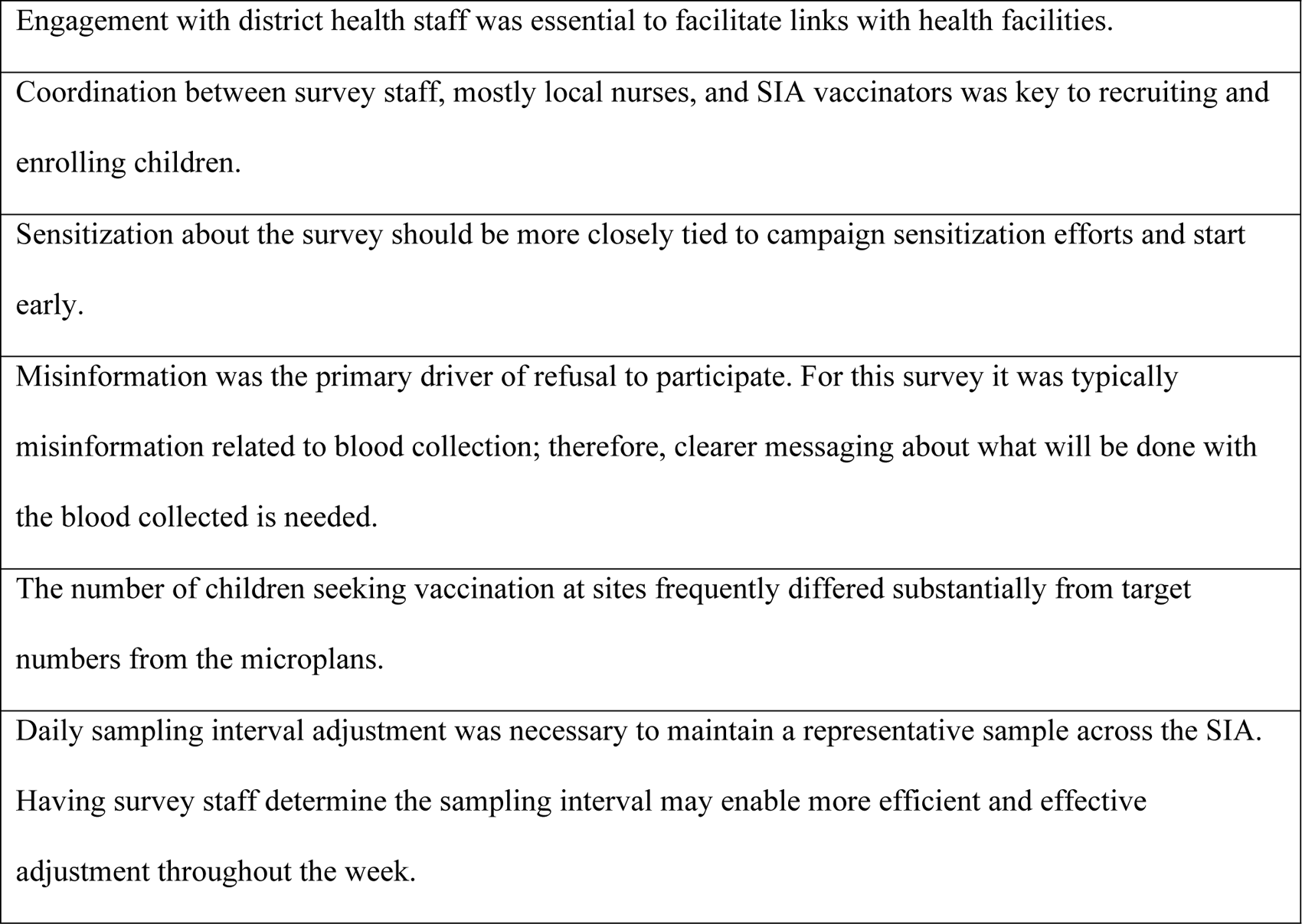
Key findings and lessons learned on implementation.

Another key aspect was to ensure sufficient social mobilization not only for the vaccination campaign to ensure children attend, but also for the blood collection. Because one of the biggest concerns for collection of blood during the campaign was that it had the potential to negatively impact the campaign, blood was collected after the child had already received their vaccine and misconceptions and refusals were closely tracked. There were no reports of parents refusing to vaccinate their children because blood might be collected from them. According to the post-campaign coverage survey, vaccination coverage for the campaign in Copperbelt Province, where Ndola District is located, was 79%, higher than the national coverage of 68%. Southern Province, where Choma District is located, was comparable to the national average (65%) (15).

While there were misconceptions about the reasons for blood collection, staff believed these could be overcome with dissemination of more information. The reasons for confusion about blood collection could have been due to incorrect explanations provided by staff. The concept that blood being taken during the campaign would provide seroprevalence estimates from before vaccination seemed difficult to explain, with only one data collector providing a correct interpretation. Additionally, the concerns about COVID testing could have resulted from the questionnaire asking about COVID vaccine acceptance (16). It was recommended that social mobilization for the survey be done far in advance, include all staff at the health facility rather than just those involved in the research, and be paired with mobilization about the campaign. Combined messaging of the research with the campaign could capitalize on social mobilization resources and avoid confusion in the community about separate planned activities occurring simultaneously.

Although it created logistical challenges, changing the sampling interval allowed us to enroll across all 6 days of the campaign and throughout the entire day. We relied on the district microplans for a baseline estimate of the number of children to be vaccinated at each survey location. In our experience, accuracy of the microplans varied by health facility, as we observed both over- and underestimates. Issues with microplans have been demonstrated for other campaigns in other settings (17). Earlier and more frequent engagement with the district health staff involved in planning the SIA may have helped to provide additional clarity on the microplans.

Although estimates from the microplans were used to inform initial sampling intervals, survey coordinators and staff worked closely during the week to adjust the sampling interval based on observed SIA volume and community settings (e.g., weather, weekly RI). However, this required survey staff to quickly modify their sampling procedures each day, which was confusing for staff and led to some situations where the changes were not implemented correctly or not at all. In a few situations the survey staff hit their maximum by midday, so afternoon SIA attendees were excluded from sampling, or the bulk of children enrolled at a site were from the first few days of the SIA. In the future we recommend local survey staff have more ownership over sampling intervals, with guidance for when and how to adjust the sampling interval and feedback loops to the survey coordinators.

Some of the limitations of this analysis are that we interviewed serosurvey and vaccination campaign staff, but we did not directly interview caregivers about the reasons for their refusals. Refusals, particularly at certain sites in Choma District, may have impacted representativeness of our study population. We acknowledge that high enrollment in the serosurvey may be due to targeting a health seeking population bringing their children for a health campaign. Therefore, results obtained from this sampling design may not be generalizable to participants who do not attend health campaigns. While we did not find evidence of the blood collection affecting vaccination campaign uptake, there was lower vaccination coverage for the 2020 November SIA compared to 2016. However, this was seen nationally and believed to be due to the COVID-19 pandemic (15).

Because this was the first experience conducting a serosurvey during a vaccination campaign, it was important to evaluate the implementation. Using a program evaluation perspective, we were able to ascertain the successes, challenges, and lessons learned of collecting dried blood spots from children attending an integrated Child Health Week (Table 1). We believe this platform provides an opportunity to integrate other services and research questions amongst children attending the campaign and can be implemented without disrupting vaccination efforts.

## Acknowledgments

We would like to acknowledge all the children and their parents who agreed to take part in this study. We would also like to acknowledge the Ministry of Health and all the serosurvey staff, vaccinators, nurse in-charges, and district health staff who contributed to the successful implementation of the project. Finally, we acknowledge Monica Pilewskie for her support coding the transcripts from the interviews.

## Data availability

The individual survey data were collected under data sharing agreements from Zambia Ministry of Health and the Zambia National Health Research Authority. As per the Zambia Health Research Act, access to data requires approval from the Zambian National Health Regulatory Authority. To obtain this access, please contact Dr Victor Chalwe, Acting Director of the Zambia National Health Research Authority.

## Supporting information captions

**S1 Table. Characteristics of staff participating in qualitative interviews.** In recruitment of participants, we tried to ensure a mix of fixed and outreach vaccination sites were represented, no overlap in team members (both team members from the same study team not included) and included study team staff and vaccination campaign team staff working at the same health facility.

**S1 Fig. Number of children enrolled by campaign day and site**

**S2 Fig. Sampling interval by campaign day and site**. Data on the sampling interval was missing at some timepoints.

**S1 Appendix. Supplemental methods**

